# Vaccination strategies for minimizing loss of life in Covid-19 in a Europe lacking vaccines

**DOI:** 10.1101/2021.01.29.21250747

**Authors:** Patrick Hunziker

## Abstract

**Aim and Background:** **We aimed at identifying vaccination strategies that minimize loss of life in the Covid-19 pandemic in a Europe lacking vaccines.** Covid-19 mainly kills the elderly, but the pandemic is driven by social contacts that are more frequent in the young. Vaccines elicit stronger immune responses in younger persons. As vaccine production is a bottleneck, many countries have adopted a strategy of first vaccinating the elderly and vulnerable, while postponing vaccination of the young.

**Methods:** Based on published age-stratified immunogenicity data of the Moderna mRNA-1273 vaccine, we compared the established “one dose fits all” approach with tailored strategies by epidemic modeling: The known differential immunogenicity of vaccine doses in different age groups is exploited to vaccinate the elderly at full dose, while the young receive a reduced dose, increasing the number of individuals receiving the vaccine early. A modeling approach at European Union scale with population structure, Covid-19 case and death rates according to Europe in late January 2021 is used.

**Results:** When the elderly were vaccinated preferentially, the pandemic initially continued essentially unchecked, as it was dominantly driven by social contacts in other age groups. Tailored strategies, including regular dosing in the elderly but reduced dose vaccination in the young, multiplied early vaccination counts, and even with some loss in protection degree for the individual person, the protective effect towards stopping the pandemic and protecting lives was enhanced, even for the elderly. In the European Union, pandemic duration (threshold >100’000 cases/day) was shortened from 53 to 18-24 days; cumulative death count over 100 days was reduced by >30’000. Data suggest that the findings may be relevant to both, the Moderna and the Pfizer-BioNTech mRNA vaccines.

**Conclusion:** Protecting the vulnerable, minimizing overall deaths and stopping the pandemic in Europe is best achieved by an adaptive vaccination strategy using an age-tailored vaccine dose.

## Introduction

Faced with the Covid-19 pandemic, vaccines against SARS-Cov2 have been developed in unprecedented speed, and mRNA vaccines like the Pfizer BNT162b2 vaccine[^1^](Tozinameran) and the Moderna mRNA-1273 vaccine[^2^] have shown excellent immunogenicity, safety and protection against disease, and data indicating that they protect against virus transmission are accumulating. While vaccines development was rapid, vaccine production capacities are now the key bottleneck for deployment. Despite its comparably high proportion of elderly and thus, vulnerable persons, Europe trails in vaccination due to slow vaccine supply.

During development, dose optimization of the Moderna vaccine has been performed towards optimal protection of the elderly, exploring doses of 25, 50, 100 and 250ug, as the elderly show reduced immune response on vacccination[^3^]. Laboratory-assessed immunity levels typically exceeded those seen in the plasma of reconvalescent patients who have a protection of 83% for at least 5 months[^4^]. Immunogenicity in the young is even higher than in the elderly. Certain Covid-19 vaccination study populations have 76% protection against infection for at least four months[^5^] despite some decline of the measured immunity parameter in the elderly, indicating that vaccination can protect against infection as well as against symptomatic disease, rendering vaccination a rational approach for stopping the pandemic. Emerging data indicate that coronavirus vaccines are blocking transmission[^6,7^], not only avoiding severe cases. We noted that in the young, a 25µg dose of the Moderna vaccine elicited an immune response level at day 57 that was comparable to the immune response seen in patients older than 71 years at day 119 (Table), a group in which the vaccine achieves >86% protection. Likewise, preprint data for the Pfizer Tozinameran vaccine show strong immunogenicity[^8^] at threefold reduced dose in the young. The interpretation that good immunogenicity translates into good protection is very plausible[^9,10^], implying that in the young, having a stronger immune response, a lower vaccine dose may suffice to achieve sufficient protection. As in mRNA lipid nanoparticle-based vaccines, dose reduction and requirement reduction for essential excipients (e.g., lipids) goes hand in hand, a dose reduction directly translates to an increased number of doses.

**Table.** Reported immune responses elicited by the Moderna vaccine. Note that the 25µg dose in the <55 year old elicits a similar or stronger immune response compared to the >71 years old at 119 days, and compares favorably with the immune levels found in convalescent plasma. (Immune titers for the young treated with 25 µg at 119 days are not available). S-2P is the antigen encoded by the vaccine. RBD ELISA measures receptor-binding domain binding antibodies. PsVNA_50_ is the pseudovirus neutralization assay’s 50% inhibitory dilution. PRNT_80_ is the live-virus plaque-reduction neutralization testing assay’s 80% inhibitory dilution.

|  | **>71y**<br>[Widge,<br>NEJM 2021] | **<55y**<br>[Jackson, NEJM<br>2020] | **Convalescent**<br>**plasma** [Jackson,<br>NEJM 2020] |
| --- | --- | --- | --- |
|  | **2x 100µg,**<br>*day 119* | **2x25µg,**<br>*day 57* |  |
| **S-2P ELISA** |  | 299'751 | 142'140 |
| **RBD ELISA** | 157'964 | 183'652 | 37'857 |
| **PsVNA<sub>50</sub>** | 109 | 80·7 | 109·2 |
| **PRNT<sub>80</sub>** | 165 | 339·7 (d43) | 158·3 |

We therefore hypothesized that exploiting age-tailored vaccination dosing may allow increasing the number of vaccinated persons in Europe faster and thereby may lead to improved pandemic control.

## Methods

The pandemic was modeled using a discrete-time[^11,12^], extended model inspired by the SEIR (“susceptible-exposed-infected-recovered”) approach with daily stepping, as shown in Figure 1 and described in detail in the supplement. Essentially, it computes daily infection rates, transmission rates (infected persons can transmit in a time window after infection), death rates (occurring with a time delay to infection) based on the social interaction between infected, nonimmune and immune persons, whereby persons after prior infection or vaccination show a reduced infection risk. The model incorporates separate, interacting age groups with age-specific immune levels triggered by vaccination or natural infection, and age-specific fatality rates. The model was initialized with population size and age structure according to the European Union with a population of 447’706’200, split into a cohort of 90’436’652 “old” persons >64 years, and 357’269’547 “young” persons ≤64, as reported in the EU Eurostat respository[^13^]. European Covid-19 case numbers were from the Johns Hopkins University CSSE dataset and were used to initialize the model to 195’000 per day as per mid-January 2021. The infectious window after Covid-19 infection was assumed to be day 1 to day 7 after infection. The model was run for 100 days.

**Figure 1:**
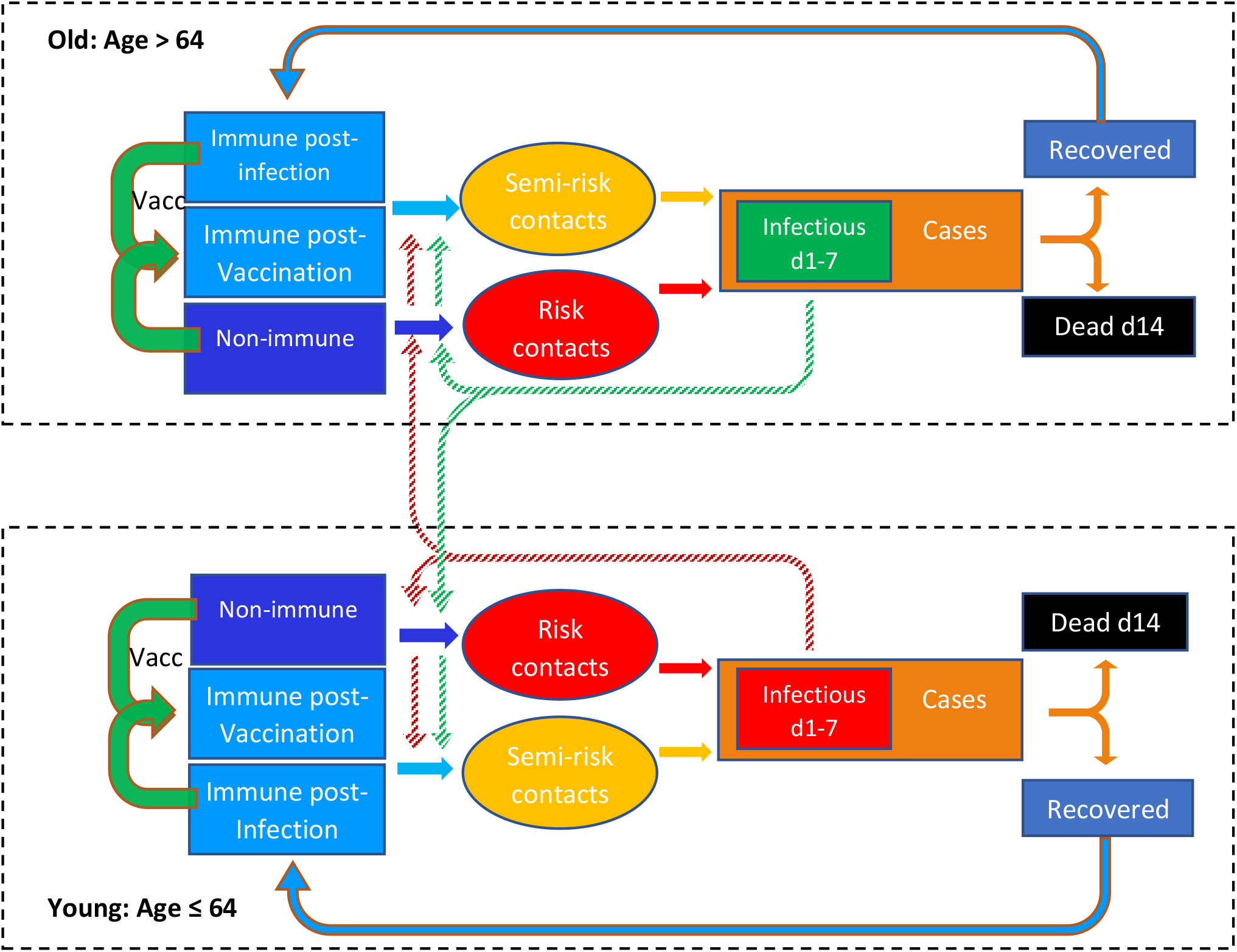
An extended, time-discrete version of the susceptible-exposed-infectious-recovered (SEIR) modeling approach. It incorporates two age strata and a tailorable degree of susceptibility in infection-naïve, vaccinated, and post-infection individuals (including silent infections). Protective efficacy of vaccination can be adapted to the age group and the vaccine dose, and vaccination campaigns can be modeled by daily setting the number of new vaccinations. Time lag and time window of infectivity after a risk contact is incorporated, and the time lag of deaths relative to diagnosis of new cases is implemented. Social interaction within and between the age strata with cross-infection uses information on vaccination count and degree of susceptibility in each group. As protection by vaccination or prior infection is not 100%, also the “semi-risk contacts” contribute to new cases. “Cases” rather than “infections” are used because this better allows parameterization of the model with real-world data; in addition, sensitivity testing has been performed by adding nondiagnosed infections up to a 1:1 ratio with “cases”. d1 and d14 indicate day 1 and day 14, respectively. “Vacc” is vaccination.

Young persons were set to have 80% of their social contacts with the “young” and 20% with the “old” [^14^], while for the old, contacts to other elderly and the young were each assumed to be 50%. Daily transmissions for each age group were derived from the daily propensity of “risk contacts”, i.e., encounters of non-immune with infectious persons of either age group, plus, weighted by vaccine protection level, of “semi-risk contacts”, i.e., encounters of a vaccinated or previously infected with an infectious person. Deaths were based on the apparent case fatality rate in the Europe in December 2020/January 2021, approximately 2.5% during a quasi-steady state in case numbers implying an R number near 1.0. The age distribution of Covid-19 deaths was computed from age-dependent mortality taken from the European Center for Disease Prevention and Control situation dashboard[^15^].

Four vaccination strategies were compared:

a. “one dose fits all”, using the standard vaccine dose, and starting with the old
b. “Adaptive”: half of vaccine supply for standard dose vaccination of the old, plus half of the vaccine supply for vaccination of the young at quarter dose (thereby reaching more young persons early)
c. “quarter dose vaccination, only to the young”, a strategy that maximizes the number of vaccinations in the young, i. e. the “frequent transmitters”, allowing for potential reduced vaccine efficacy in the individual person.
d. Standard dose, starting with the young

Vaccine stock is available for 1 million full dose vaccinations (Moderna, 2×100 µg) per day. Vaccination efficacy was parameterized with data reported by Moderna on phase I[^16,17,18,19,20^] II and III[^21^] studies. The interval from vaccination to protection was 10 days. Protective efficacy for the 100µg vaccine dose was 95.6% in the young and 86.2% in the elderly as reported; in one analysis, the vaccine efficacy for avoidance of transmission of a 25µg dose in the young was set to 86.2% based on the levels of immunogenicity achieved in the young compared to the immune response in the elderly vaccinated with 100µg; then, transmission blocking efficacies were varied between 30% and 90% for the 25µg dose in the young to explore the impact of blocking efficacy on strategy preference.

Further sensitivity testing was performed for the number of available vaccines, the proportion of unrecognized infections, and variations in the infectiosity of the virus strain, as described in more detail in the supplemental material.

### Ethics

According to a written statement from the relevant Ethical Kommittee Nordwestschweiz EKNW, computer modeling studies not including subjects do not fall under their ethical committee jurisdiction.

## Results

### Results are summarized in Figure 2

A**“one dose fits all” vaccination strategy** starting with the elderly allows an initially unchecked propagation of the virus in younger population segments with high “risk contact” numbers, as shown in figure 2, panel A; a large number of infections in the young inevitably spreads to some degree to the elderly (who are not 100% protected by the vaccine), with case numbers above 100’000 per day up to day 53, 199’000 cumulative deaths over 100 days and death rates of >1000 deaths per day until day 80.

**Figure 2:**
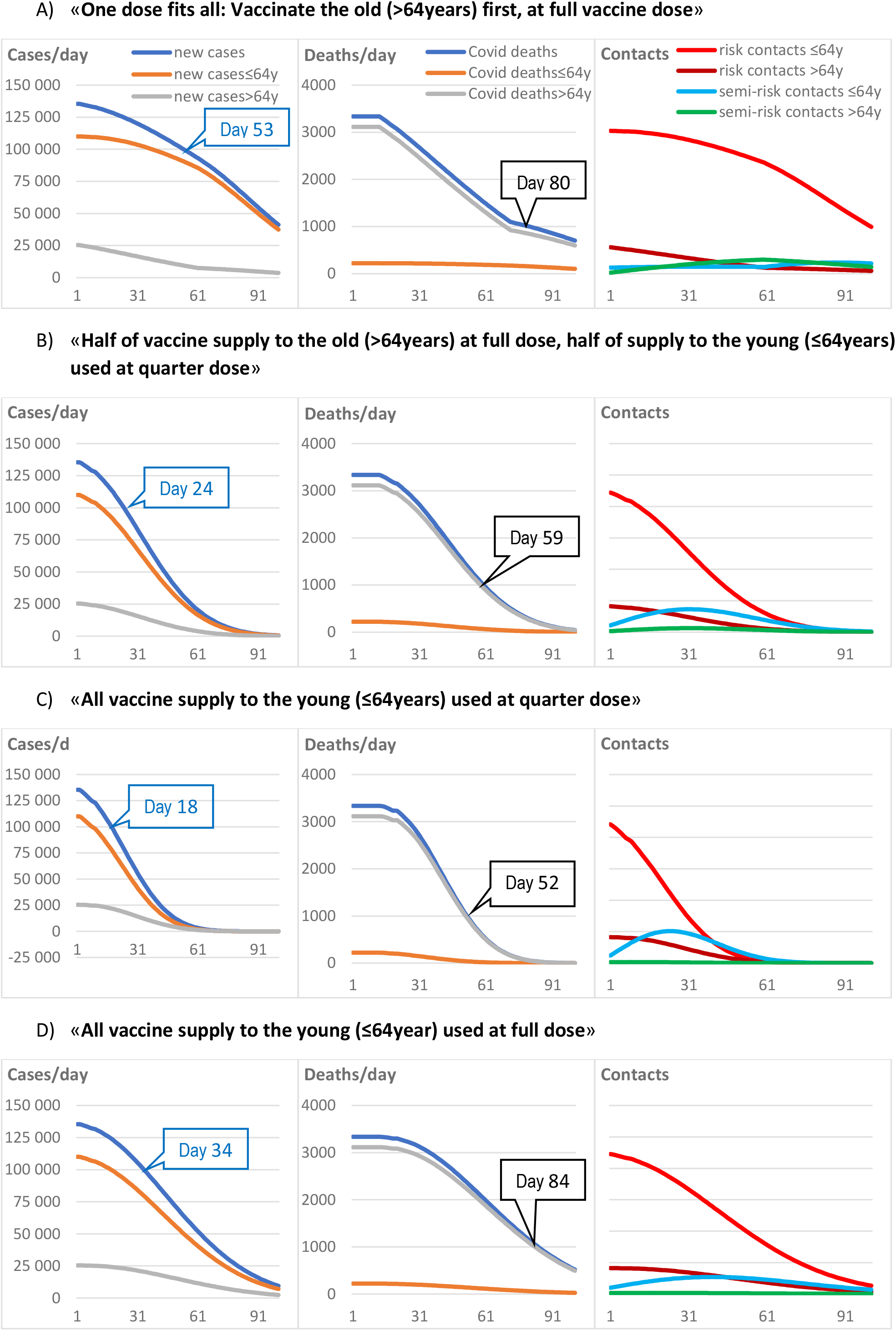
Impact of different vaccination strategies on Covid-19 cases, deaths and the propensity of “risk contacts”. Panel A) vaccinating the elderly first leaves large population segments unprotected. Note that the prolonged spread of infection in the non-vaccinated young spills over to high case and death counts in the elderly that are not yet vaccinated or only partly protected by the vaccine. Panel B) using half of the vaccine supply to vaccinate the elderly at full dose, and using the other half of the supply to vaccinate the young with a quarter dose, leads to a faster decline in cases and fewer deaths, even in the elderly. Panel C) Using all vaccines for vaccinating the young with a quarter dose leads to faster reduction of overall case numbers, and thereby, indirectly, also protect the elderly after a sufficient overall vaccination rate is achieved, yielding low overall deaths. Panel D) Compared to the best scenarios, using the full vaccine dose for vaccinating only the young (“the frequent transmitters”) is slower compared to the quarter dose strategies in stopping the pandemic despite being more protective for the individual person, and is associated with substantial death counts in the elderly. “Risk contact” designates an encounter of a non-immune with an infectious person, and “semi-risk contact” designates an encounter of a vaccinated person with an infectious person, taking into account that vaccine protection is less than 100%. X-axes are days. Blue labels show the day when case numbers fall below 100′000. Black labels show the day death numbers fall below 1000.

In contrast, a strategy initially **vaccinating the young only**, at full vaccine dose, shown in panel D, stops the pandemic earlier with case numbers falling below 100’000 on day 34, but lacking protection for the vulnerable leads to 226’000 deaths, with death rates falling below 1000 on day 84.

In contrast, an **adaptive strategy** using half the available vaccine stock for vaccinating the elderly at full vaccine dose and using the other half at a reduced dose (figure 2, panel B) to immunize a much larger number of younger people, even at the price of a somewhat reduced vaccine efficacy per individual person, is much more effective in reducing case numbers and deaths in each age group. Using a quarter dose for the young (25µg), assuming 86.4% protection, allows shortening the time to <100’000 cases to 24 days and reducing deaths to 165’000 at 100 days. This scenario also reaches the milestone of <1000 deaths per day significantly faster, in 59 days.

Distributing a **quarter dose (25µg) only to the young** (i.e., not vaccinating the elderly; figure 2, panel C), also assuming 86.4% protection, reduced deaths to 148’000, protecting the elderly indirectly by shortening the pandemic, with< 100’000 cases per day reached on day 18 and <1000 deaths per day reached on day 52.

Results proved to be robust against varying input parameters, including variation of the transmission blocking efficacy of a vaccine: The adaptive strategy combining quarter dose vaccination in the young and full dosing in the old remained preferable to the “one dose fits all” approach starting with the old, down to a transmission blocking vaccine efficacy of 30% (Figure 3 A). Sensitivity testing for proportion of unrecognized infections from 0 up to 1:1 relative to the confirmed case numbers did not alter the benefit of the adaptive strategy compared to the “elderly first, one dose fits all” strategy, but slightly reduced case numbers in all scenarios (Figure 3B). Availability of vaccine doses within 0.5-2.5 million full vaccinations per day preserved the advantage of the adaptive vaccination strategy (Figure 3C).

**Figure 3:**
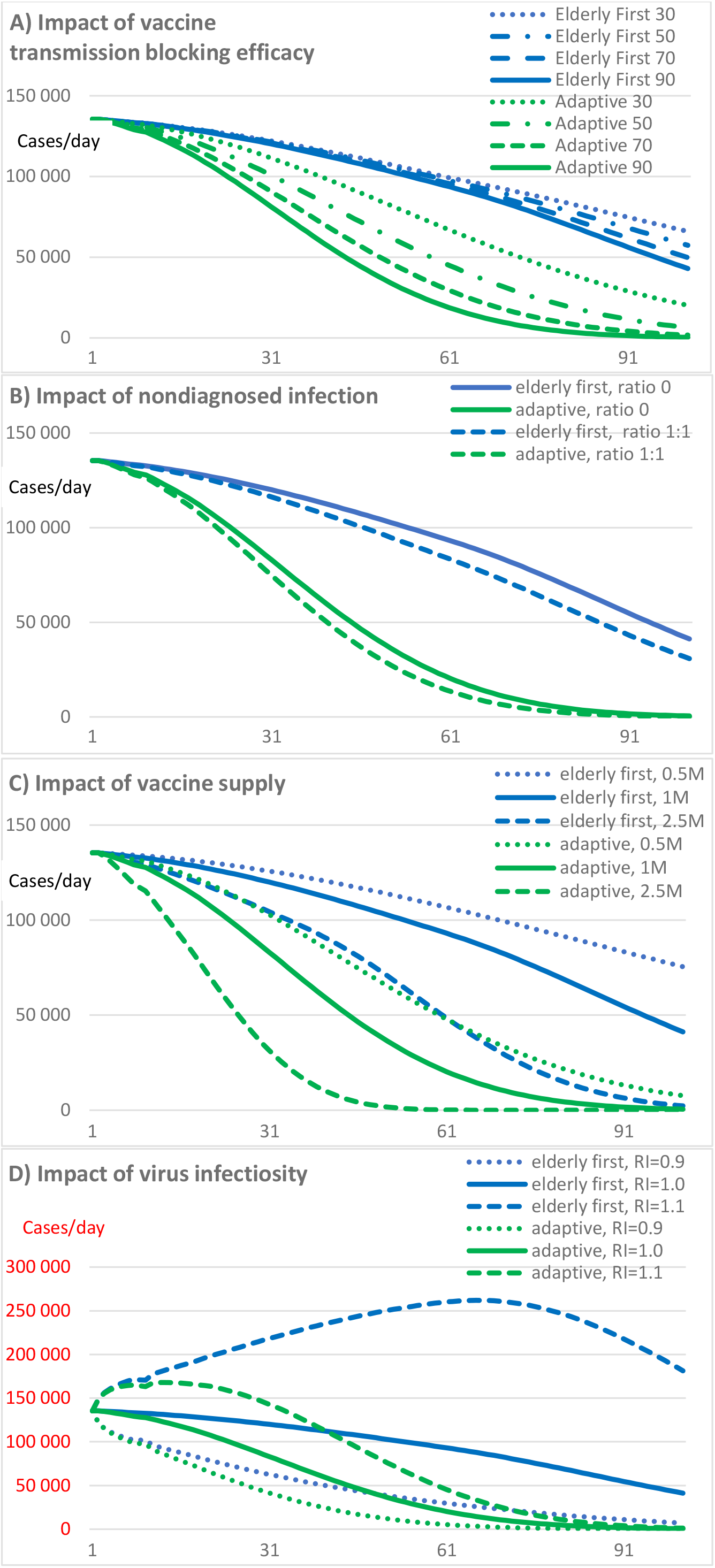
Panel A) Sensitivity of choice of strategy to different efficacies for transmission blocking or vaccine- and infection-mediated immunity. Transmission blocking efficacy for vaccine and infection-mediated immunity is varied between 30 and 90% for the young. For the old, a transmission blocking efficacy is kept at 86.4%, potentially biasing the results in favor of the “one dose fits all” strategy starting with the old. Nevertheless, the adaptive strategy, reaching large numbers of young persons early, is more effective in reducing the Covid-19 case load at all transmission blocking efficacies tested, down to 30%. Numbers in the label indicate transmission blocking efficacy of the vaccine in percent. Y-axis is cases/day. Panel B) Information about nondiagnosed infections in Europe are still sparse but seroprevalence is far from a herd immunity prevalence. In this context, the model indicates that including a ratio of nondiagnosed infections from 0 to 1:1 (i.e., equal number of diagnosed and nondiagnosed cases) only has a limited impact on the course of the epidemic and the benefit of an adaptive vaccination strategy is maintained. Panel C) At each level of vaccine supply, the adaptive strategy outperforms the conventional one. Panel D) The adaptive strategy is particularly beneficial in a situation where the epidemiologic reproduction number is >1.0 due to spread of a more infectious virus strain, as currently observed in many European countries. Note the different Y scale compared to panels A-C.

**Changes in infectiosity of the dominant virus variant** have a major impact on case numbers, as shown in figure 3D. While the baseline scenario starts with an epidemiologic reproduction number of 1.0, spread of a virus variant with increased infectiosity that is not fully contained by societal measures and therefore results in an initial reproduction number of 1.1 is followed by a next infection wave. In this scenario (as currently observed in many European countries), the benefit of the adaptive strategy to reduce case numbers is markedly larger, compared to the conventional vaccination strategy. In contrast, if strict societal measures lead to a decrease a low epidemiologic reproduction number to 0.9, as potentially achievable with strict lockdown, case counts and deaths are reduced as expected, although the adaptive strategy maintains a benefit.

## Discussion

Combining demographic and epidemiologic data from Europe with published immune responses to mRNA vaccines, we find that vaccination strategies tailored to the characteristics of vaccines and its recipients have a large potential for saving lives and shortening the pandemic, compared to the widely used but slow “one dose fits all” and “elderly first” approach. Specifically, the strong, age-dependent immunogenicity of the Moderna vaccine that shows stronger immune responses in the young but still has excellent efficacy at full dose in the elderly inspires an adaptive strategy that combines full dose vaccination in the elderly with fractional dose vaccination in the socially more active, young[^22^] to reach a much larger number of persons with the vaccine, faster. The resulting acceleration of vaccinations, the reduction in cases and deaths, and the shortening the pandemic promise to better preserve healthcare systems and economy, and may also accelerate access to vaccines for poorer countries in a time when nations risk to quarrel over this resource. Notably, “protecting the vulnerable” is achieved best, when vaccination not only focuses on the vulnerable, but from early on includes the young drivers of the pandemic[^23,24,25^]. A large number of infected young people combined with the imperfect protection achievable by vaccination still represents a relevant threat to the vulnerable and calls for a more comprehensive approach. Notably, early in a wave of infections with a strain of higher infectiosity as currently observed in Europe, an adaptive strategy was particularly beneficial.

The effectiveness of the Moderna vaccine even before the maximal immune response has been achieved is evident already 10 days after the first dose in the pivotal trial. Emerging data confirm a protective effect of mRNA Covid-19 vaccines for blocking transmission[^26^], even in the elderly, and before the full immune response is reached[^27,28^]. Protection against reinfection after natural infection[^29^] despite lower immune titers and experience with “fractional” dose vaccination in other viral diseases[^30,31^] further support this concept. The societal benefit from reduced-dose vaccination results from a non-linear vaccine dose-efficacy relationship, where the fractional loss of individual protection is less than the gain in vaccine doses. We note that in the Pfizer Tozinameran (BNT162b2) mRNA vaccine, antibody and T-cell responses show a similarly nonlinear dose-response relationship[^32^], with only a marginal reduction in immune titers when dose is reduced from 30 to 10ug, suggesting that for the Pfizer vaccine too, a 3-fold dose reduction and thus a 3-fold increase in early vaccinations for the young might be feasible.

While the Moderna vaccine retains good activity for the currently spreading mutant of concern B.1.1.7[^33^], thus preserving the rationale of this strategy, future variants of concern that evade the immune response from prior infection or from first-generation vaccines are on the rise[^34^], may require additional vaccines, and will put further strain on production lines, thus rendering optimized vaccination strategies even more desirable. The risk of mutants developing *because of* reduced dose vaccination may be limited[^35^] because virus infection after coronavirus vaccination typically induces a strong immunity boost[^36^].

Study limitations include the interpretation and extrapolation of published immunogenicity data that are solid but still limited; such analyses would be further strengthened by the availability of more, age-dependent immunity parameters over time after vaccination, in particular for various doses at longer term. Using laboratory surrogate parameters for protective vaccine efficacy is increasingly accepted[^37,38^], and vaccine safety has amply been documented. Likewise, evidence for vaccine transmission blocking efficacy is recent but appears consistent; the model was robust against large variations in this potential confounder. Epidemic models are always a simplification of the spatiotemporal and biological disease complexity and evolution; nevertheless, our findings were robust for several possible confounders. As we focus on high-efficacy mRNA vaccines here, generalizations to other vaccines will require examining their specific dose-response relationships. Pivotal studies for this vaccination approach are easily implementable: Allocate entire cities to an adaptive strategy, and count cases, deaths and outbreak duration. Applying this “off-label” use of a registered drug in patients will require specific ethical and regulatory permits.

## Conclusion

Adaptive vaccination strategies, namely fully dosing a vaccine in the elderly, vulnerable, and concomitantly applying a highly effective mRNA vaccine at reduced dose in the remainder of the European population, or even a reduced dose strategy focused on the young only, will multiply the number of persons receiving the vaccine early, may contribute to stopping the pandemic faster and have the potential to save many lives. Evidently, the vulnerable are best protected by protecting society as a whole.

## Data Availability

A spreadsheet with daily data will be available, starting with publication in a scientific journal, for 6 months, by email request from the author, for non-commercial, scientific purposes by academic institutions and government agencies, with mandatory source attribution when used. For use of the statistical model and for other uses, contact the author directly.

## Data sharing statement

A spreadsheet with daily data will be available as supplementary material, for non-commercial, scientific purposes by academic institutions and government agencies, with mandatory source attribution when used. For use of the statistical model and for other uses, contact the author directly.

## Research in Context

### Evidence before this study

As source of demographics of Europe, we used the official EU Eurostat repository. For epidemiology of the Covid-19 pandemic, the Johns Hopkins University CSSE dataset and the European Center for Disease Prevention and Control situation dashboard were used. For immunogenicity of the Moderna vaccine, PubMed was used to identify the published data related to “mRNA-1273”; a Bing search retrieved an additional presentation slide set from Moderna about its testing pipeline. Vaccine supply limitations are amply referred to in official statements published by the media outlets. Efficacy for the Moderna vaccine at regular dose is 95.6% (95% CI, 90.6-97.9) for those up to 64 years and 86.4% (95% CI, 61.4-95.2%) for those above, as reported in the pivotal phase 3 publication. Immunogenicity data are given in the table.

### Added value of this study

Using an epidemic model initialized with EU-wide population, Covid-19 case and death data from mid-January 2021, alternative, up to now unexplored vaccination strategies were defined and compared to the currently preferred approach that consists of initially focusing vaccination on the elderly because vaccine supplies are insufficient for a broader initial use. The study addressed several alternative vaccination strategy scenarios, in particular the use of a reduced vaccine dose in the group that showed the strongest immune response to vaccination, namely those < 65 years.

### Implications of all the available evidence

Vaccination, combined with societal measures up to lock down, will be the mainstay of mastering the SARS-CoV2 pandemic. The available evidence, including the findings reported here, imply that tailored vaccination schemes adapted to the specific characteristics of the vaccine, the demographics and the immune response of population subgroups have a large potential for reducing case numbers and deaths in Europe.

## Supplementary materials

### Model construction

The model is inspired by the SEIR approach but is performed in discrete-time steps and therefore uses difference equations instead of continuous time and differential equations. This approach allows parameterizing the model in a straightforward way by daily statistical case, death and vaccination counts available from the various data sources. The model has discrete structure with a time step of one day for 100 consecutive days.

Two age strata are modeled, allowing to use age-group adapted immunity, fatality, and social interaction propensity. The model is initialized with the population size of the European Union split in two age segments, ≤ 64 years (“young”) and 64 years(“old”), using the daily infection rate and the cumulative number of cases documented in mid-January 2021.

In each age stratum, an infection-naïve, a vaccinated and a postinfective group is modeled, with transitions between groups upon infection or vaccination. Susceptibility to transition to “infection” is 1.0 for the infection-naïve group and parameterizable for the other groups to allow for modeling of various vaccine efficacies.

Contact modelling between persons assumes that a “young” person has 80% of its social encounters with the “young” and 20% with the “old”, while “old” persons have 50% of its social encounters with the “old”. “Risk contacts” are defined as the proportion of encounters (relative to day 1) of noninfected persons with persons newly infected within the past 7 days, and “Semi-Risk contacts” are defined as the proportion of encounters of immune (natural or post vaccination) persons with newly infected persons, using separate computation for each age group. Daily contact frequency is conservatively chosen as equal in both groups. Selecting larger contact numbers in the young would further underline the key findings of the study.

During December 2020/January 2021, number of cases, deaths and tests stagnated at a high level in Europe, implying an approximate overall R value of 1.0 and allowing to estimate an approximate case fatality rate during steady state conditions, and to initialize an initial contact propensity by multiplying the number of infectious persons with susceptible persons for each stratum (weighted for the degree of immunity). We couple the pandemic modeling to the documented (though imprecise) case count rather than to “infection” count because testing rates are currently high and true infection rates (including asymptomatic individuals) across Europe are currently at best very coarse estimates. To assess the impact of nondiagnosed infections that will also make persons transition from “naïve” to “post-infection” (a cause of bias that is minor early in a pandemic but may become more important when large proportions of a population have been infected), sensitivity testing for such unrecognized in infections was performed by test runs with added proportions of nondiagnosed infections in a range from 0 to 1:1 (i.e., same number of nondiagnosed infections as diagnosed cases) and found that including such an unknown number of nondiagnosed infection does not alter the key findings of the study.

The number of new cases for is computed as proportional to the number of risk contacts plus the semi-risk contacts scaled by the degree of immunity (i.e., effectivity of the vaccination for this group, or natural immunity). The infectious window is on day 1 to 7 after infection.

The number of deaths was modeled as proportional to the number of new cases, using the case fatality rate as described in the methods section, with a time lag of deaths after infection of 14 days.

1 million vaccine full vaccine sets (2*100 µg) per day are supplied. In the baseline “elderly first” scenarios, they are either applied as 1 million full vaccinations of the >64 year old person per day in the standard vaccination strategy until 65% of the elderly population is vaccinated, followed by 1 million full vaccinations per day in the young. In the the adaptive scenario, daily 500’000 vaccinations are performed in the elderly and 2 million vaccinations at quarter dose in the young. In the other scenarios, 4 million vaccinations at quarter dose limited to the young are performed, or 1 million vaccinations at full dose are given exclusively to the young, respectively. Vaccinated persons became protected against infection on day 10, to a degree corresponding to the level of immunity conferred by the given vaccine dose and age group. Sensitivity testing varying the supply of available vaccines from 0.5 to 2.5 million full vaccinations has been performed; while decreasing the number of vaccines slows the overall progress of the vaccination campaign and better supply speeds it up, the advantages of the adaptive strategy and the main conclusions of the study remain unchanged.

Counts of population, newly infected, infectious, immune, susceptible, vaccinated persons and deaths were updated daily.

GATHER checklist of information that should be included in reports of global health estimates

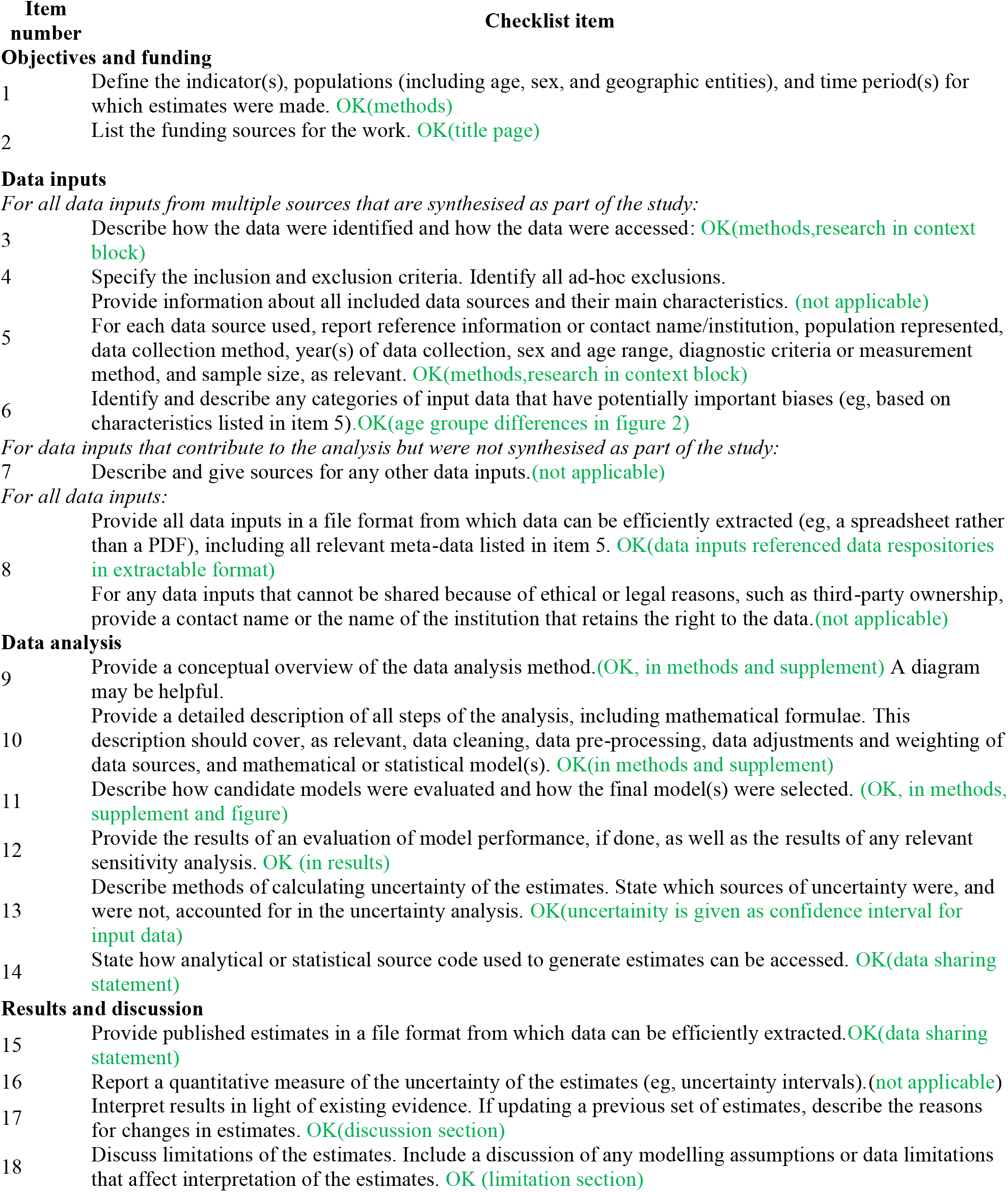

## References

1 Polack F P, Thomas S J, Kitchin N, et al. Safety and efficacy of the BNT162b2 mRNA Covid-19 vaccine N Engl J Med 2020; 383, 2603–15.

2 Baden LR, El Sahly HM, Essink B, et al. Efficacy and safety of the mRNA-1273 SARS-CoV-2 vaccine N Engl J Med December 30, 2020. DOI: 10.1056/NEJMoa2035389.

3 Varnica Bajaj V, Nirupa Gadi N, Allison P. Spihlman AP et al. Aging, Immunity, and COVID-19: How Age Influences the Host Immune Response to Coronavirus Infections? Front. Physiol., 2021. DOI : doi.org/10.3389/fphys.2020.571416

4 Lumley S F, O’Donnell D, Stoesser NE, et al. Antibody status and incidence of SARS-CoV-2 infection in health care workers. N Engl J Med 2020; DOI: 10.1056/NEJMoa2034545

5 Voysey M, Costa Clemens SA, Madhi SA et al., Single dose administration, and the influence of the timing of the booster dose on immunogenicity and efficacy of ChAdOx1 nCoV-19 (AZD1222) vaccine. Preprint, Available at SSRN: https://ssrn.com/abstract=3777268

6 Dvir Aran. Estimating real-world COVID-19 vaccine effectiveness in Israel using aggregated counts. medRxiv 2021; doi: https://doi.org/10.1101/2021.02.05.21251139

7 Voysey M, Costa Clemens SA, Madhi SA et al. Single-dose administration and the influence of the timing of the booster dose on immunogenicity and efficacy of ChAdOx1 nCoV-19 (AZD1222) vaccine: a pooled analysis of four randomised trials. The Lancet 2021; DOI:https://doi.org/10.1016/S0140-6736(21)00432-3

8 Sahin U, Muik A, Vogler I, et al. BNT162b2 induces SARS-CoV-2-neutralising antibodies and T cells in humans. medRxiv. 2020; DOI: doi.org/10.1101/2020.12.09.20245175.

9 Lumley S F, O’Donnell D, Stoesser NE, al. Antibodies to SARS-CoV-2 are associated with protection against reinfection. medRxiv 2020; DOI: 10.1101/2020.11.18.20234369

10 Kim JH, Marks F, Clemens JD. Looking beyond COVID-19 vaccine phase 3 trials. Nature Medicine. 2021 Jan 19:1–7. 11

11 Ibeas, A., de la Sen, M., Alonso-Quesada, S. et al. Stability analysis and observer design for discrete-time SEIR epidemic models. Adv Differ Equ 2015 ; 2015, 122. https://doi.org/10.1186/s13662-015-0459-x

12 Hu, Z, Teng, Z, Jiang, H: Stability analysis in a class of discrete SIRS epidemic models. Nonlinear Anal., Real World Appl. 2012; 13, 2017–33

13 https://ec.europa.eu/eurostat/databrowser/view/tps00010/default/table?lang=en last accessed on January 25, 2021

14 Wallinga J, Teunis P, Kretzschmar M. Using data on social contacts to estimate age-specific transmission parameters for respiratory-spread infectious agents. American Journal of Epidemioogyl. 2006; 164:936–44.

15 https://qap.ecdc.europa.eu/public/extensions/COVID-19/COVID-19.html#enhanced-surveillance-tab last accessed on January 25,2021

16 Jackson L A, Anderson EJ, Rouphael NG, et al. An mRNA vaccine against SARS-CoV-2—preliminary report. N Engl J Med 2020; DOI: 10.1056/NEJMoa2022483.

17 Anderson E J, Rouphael NG, Widge AT, al. Safety and immunogenicity of SARS-CoV-2 mRNA-1273 vaccine in older adults. N Engl J Med N Engl J Med 2020; 383: 2427–38.

18 Widge A T, Rouphael NG, Jackson LA, et al. “Durability of responses after SARS-CoV-2 mRNA-1273 vaccination.” N Engl J Med 2021; 384: 80–2.

19 Supplement to: Jackson LA, Anderson EJ, Rouphael NG, et al. An mRNA vaccine against SARS-CoV-2 — preliminary report. N Engl J Med 2020; 383:1920–31. DOI: 10.1056/NEJMoa2022483

20 Jacqueline M. Miller. mRNA-1273 Clinical Development Program. Moderna presentation, August 26, 2020, retrieved from https://www.cdc.gov/vaccines/acip/meetings/downloads/slides-2020-08/COVID-02-Miller-508.pdf

21 Baden LR, El Sahly HM, Essink B, et al. Efficacy and safety of the mRNA-1273 SARS-CoV-2 vaccine N Engl J Med 2020. DOI: 10.1056/NEJMoa2035389.

22 Fumanelli L, Ajelli M, Manfredi P, Vespignani A Merler S. Inferring the structure of social contacts from demographic data in the analysis of infectious diseases spread. PLoS Comput Biol 2012 ; 8, Article e1002673. DOI : doi.org/10.1371/journal.pcbi.1002673

23 Matrajt L, Eaton J, Leung T, Brown ER. Vaccine optimization for COVID-19: who to vaccinate first? medRxiv 2020; DOI: 10.1101/2020.08.14.20175257

24 Meehan MT, Cocks DG, Caldwell JM, et al. Age-targeted dose allocation can halve COVID-19 vaccine requirements. medRxiv 2020; DOI: 10.1101/2020.10.08.20208108

25 Brüningk SC, Juliane Klatt J, Stange M, et al. Determinants of SARS-CoV-2 transmission to guide vaccination strategy in a city. medRxiv 2020.12.15.20248130; DOI: 10.1101/2020.12.15.20248130

26 Mallapaty S. Are COVID vaccination programmes working? Scientists seek first clues. Nature 2021 ; 589, 504–5. DOI: 10.1038/d41586-021-00140-w

27 Matan Levine-Tiefenbrun, Idan Yelin, Rachel Katz, Esma Herzel, Ziv Golan, Licita Schreiber, Tamar Wolf, Varda Nadler, Amir Ben-Tov, Jacob Kuint, Sivan Gazit, Tal Patalon, Gabriel Chodick, Roy Kishony. Decreased SARS-CoV-2 viral load following vaccination. medRxiV 2021; doi: https://doi.org/10.1101/2021.02.06.21251283

28 First indication of the effect of COVID-19 vaccinations on the course of the outbreak in Israel De-Leon H, Calderon-Margalit R, Pederiva F, Ashkenazy Y, Gazit D. medRxiv 2021; doi: https://doi.org/10.1101/2021.02.02.21250630

29 Lumley S F, O’Donnell D, Stoesser NE, et al. Antibody status and incidence of SARS-CoV-2 infection in health care workers. N Engl J Med 2020; DOI: 10.1056/NEJMoa2034545

30 Resik S, Tejeda A, Diaz M, et al. Boosting immune responses following fractional-dose inactivated poliovirus vaccine: a randomized, controlled trial. The Journal of infectious diseases 2017, 215, 175–82.

31 de Menezes Martins R, Maria de Lourdes S M, de Lima S M B Duration of post-vaccination immunity to yellow fever in volunteers eight years after a dose-response study. Vaccine 2018; 36, 4112–7.

32 Sahin U, Muik A, Vogler I, et al. BNT162b2 induces SARS-CoV-2-neutralising antibodies and T cells in humans. medRxiv. 2020; DOI: doi.org/10.1101/2020.12.09.20245175.

33 Wu K, Werner AP, Moliva JI, et al. mRNA-1273 vaccine induces neutralizing antibodies against spike mutants from global SARS-CoV-2 variants. medRxiv 2020: DOI:10.1101/2021.01.25.427948

34 Faria NR, Morales Claro I, Candido D, et al, on behalf of CADDE Genomic Network. Genomic characterisation of an emergent SARS-CoV-2 lineage in Manaus: preliminary findings. Retrieved Jan 26,2021 on from https://virological.org/t/genomic-characterisation-of-an-emergent-sars-cov-2-lineage-in-manaus-preliminary-findings/586

35 Kennedy DA, Read AF. Why the evolution of vaccine resistance is less of a concern than the evolution of drug resistance. PNAS. 2018; 115:12878–86.

36 Callow KA, Parry HF, M. Sergeant M, Tyrrell DAJ. The time course of the immune response to experimental coronavirus infection of man. Epidemiol. Infect. 1990; 105, 435–46

37 Tan CW, Chia WN, Qin X, Liu P, Chen MI, Tiu C, Hu Z, Chen VC, Young BE, Sia WR, Tan YJ. A SARS-CoV-2 surrogate virus neutralization test based on antibody-mediated blockage of ACE2–spike protein–protein interaction. Nature biotechnology. 2020;38:1073–8.

38 Jin P, Li J, Pan H, Wu Y, Zhu F. Immunological surrogate endpoints of COVID-2019 vaccines: the evidence we have versus the evidence we need. Signal Transduction and Targeted Therapy. 2021 Feb 2;6(1):1–6.

